# Transcriptional and translational genetic control in blood contributes significantly to complex trait heritability and polygenic prediction

**DOI:** 10.1101/2025.05.24.25328265

**Authors:** Shouye Liu, Xiaoyu Qian, Yang Wu, Zhili Zheng, Michael E. Goddard, Jian Yang, Peter M. Visscher, Jian Zeng

**Affiliations:** Institute for Molecular Bioscience, The University of Queensland, Brisbane, Queensland, Australia; Institute of Rare Diseases, West China Hospital of Sichuan University, Chengdu, China; Program in Medical and Population Genetics, Broad Institute of Harvard and MIT, Cambridge, Massachusetts, USA; Stanley Center for Psychiatric Research, Broad Institute of Harvard and MIT, Cambridge, Massachusetts, USA; Analytic and Translational Genetics Unit, Massachusetts General Hospital, Boston, Massachusetts, USA; Faculty of Veterinary and Agricultural Science, University of Melbourne, Parkville, Victoria, Australia; Biosciences Research Division, Department of Economic Development, Jobs, Transport and Resources, Bundoora, Victoria, Australia; School of Life Sciences, Westlake University, Hangzhou, Zhejiang, China; Westlake Laboratory of Life Sciences and Biomedicine, Hangzhou, Zhejiang, China; Nuffield Department of Population Health, University of Oxford, Oxford, United Kingdom

## Abstract

Genetic variants can influence complex traits by regulating gene products such as gene expression, RNA splicing, and protein abundance. While molecular quantitative trait loci (molQTL) are widely leveraged to infer causal genes, their overall contribution to complex trait variation remains unclear. Here, we systematically evaluate the contributions of expression (eQTL), splicing (sQTL), and protein (pQTL) QTL in blood to SNP-based heritability and polygenic prediction across 27 complex traits. Using SBayesRC, we show that these molQTL, covering only ∼1% of all SNPs, capture on average 20% of SNP-based heritability and 34% of prediction accuracy, with particularly strong contributions for blood-related traits. These estimates are comparable to those from predicted variant deleteriousness and exceed other functional annotations. Adjusting for sample size and genome coverage differences across molQTL datasets, we highlight the importance of sQTL and pQTL relative to eQTL. Our findings underscore the key role of transcriptional and translational genetic control in shaping complex trait genetic architecture.

## Introduction

The genetic control of gene products plays a central role in linking genetic variants to complex traits and diseases^1^. This is supported by the observations that most genes have regulatory variants influencing their gene expression (expression quantitative trait loci, eQTL)^2^, RNA splicing (splicing QTL, sQTL)^3^, or protein abundance (protein QTL, pQTL)^4^, and that many trait-associated variants identified from genome-wide association studies (GWAS) are located in non-coding regions enriched for these QTLs^5^. Large-scale molecular QTL (molQTL) mapping efforts, such as the eQTLGen consortium^2^, the eQTL Catalogue^6^, the UK Biobank Pharma Proteomics Project (UKB-PPP)^7^, and the Genotype-Tissue Expression (GTEx) project^3^, have created valuable resources for investigating the genetic basis of regulatory variation. These resources have been widely used to identify functionally relevant genes for complex traits by integrating molQTL data with GWAS through statistical methods such as transcriptome-wide association studies (TWAS), colocalization, and mendelian randomization^8–11^.

Although the genetic regulation of gene products presents biologically plausible mechanisms for how variants affect complex traits, recent studies have shown only limited overlap between GWAS hits and eQTL^12–14^ and only a modest proportion (∼11%) of trait variation mediated by gene expression^15^. This apparent discrepancy suggests alternative mechanisms, beyond transcriptional regulation, or context-specific genetic effects as potential explanations. Indeed, previous findings indicate that sQTL plays a crucial role in complex trait variation^16^, with an independent and comparable contribution to eQTL^17^. Further integration of multiple omics layers, including epigenetic regulation QTL, has revealed that approximately 50% of GWAS loci overlap with at least one molecular phenotype^18^. However, Hormozdiari et al^19^ reported that while stratified linkage disequilibrium score regression (sLDSC^20^) detected SNP-based heritability enrichment across molQTL datasets, sQTL were not significantly enriched when conditioning on other molQTL types. In contrast, a recent study in livestock genetics found that cis and trans eQTL and sQTL across 16 tissues jointly explain ∼70% of heritability for 37 complex traits^21^. Given these contradictory findings, the extent to which different molQTL, particularly those related to gene product regulation, contribute to trait variation remains unclear. The discrepancies observed across studies may stem from differences in sample size and genome coverage across molQTL types and the statistical methodology used for inference. To date, a comprehensive and comparative evaluation of the contributions of molQTL to complex trait variation in humans, including their role in polygenic prediction, is still lacking.

In this study, we systematically evaluated the contribution of cis molQTL involved in transcriptional and translational regulation (cis eQTL, sQTL, and pQTL) to SNP-based heritability and polygenic prediction using SBayesRC, a state-of-the-art Bayesian model that incorporates genomic annotations with GWAS summary statistics (Fig. 1; Methods). We constructed molQTL annotations incorporating eQTL^2^, sQTL^3^, and pQTL^7^ data derived from blood samples (Table 1), due to their relatively large sample sizes (compared to those from other tissues) and established relevance to immune-related and metabolic traits^22,23^. Previous research suggests that evolutionary constraints shape genetic variation in complex traits, with variants in conserved regions playing a significant role^24–26^. To benchmark the contribution of molQTL annotations, we included a binary annotation derived from the Combined Annotation Dependent Depletion (CADD) score, which incorporates conservation and other functional data to predict variant deleteriousness^27^, along with 15 core functional genomic annotations from the LDSC baseline annotation set^20^. To control for sample size discrepancies across molQTL datasets, we proposed a method for down-sampling molQTL data at the summary statistics level (Methods), enabling fair comparisons without confounding due to differences in statistical power. Our analyses were conducted across 27 complex traits and diseases using data from the UK Biobank (UKB)^28^.

**Figure 1.**
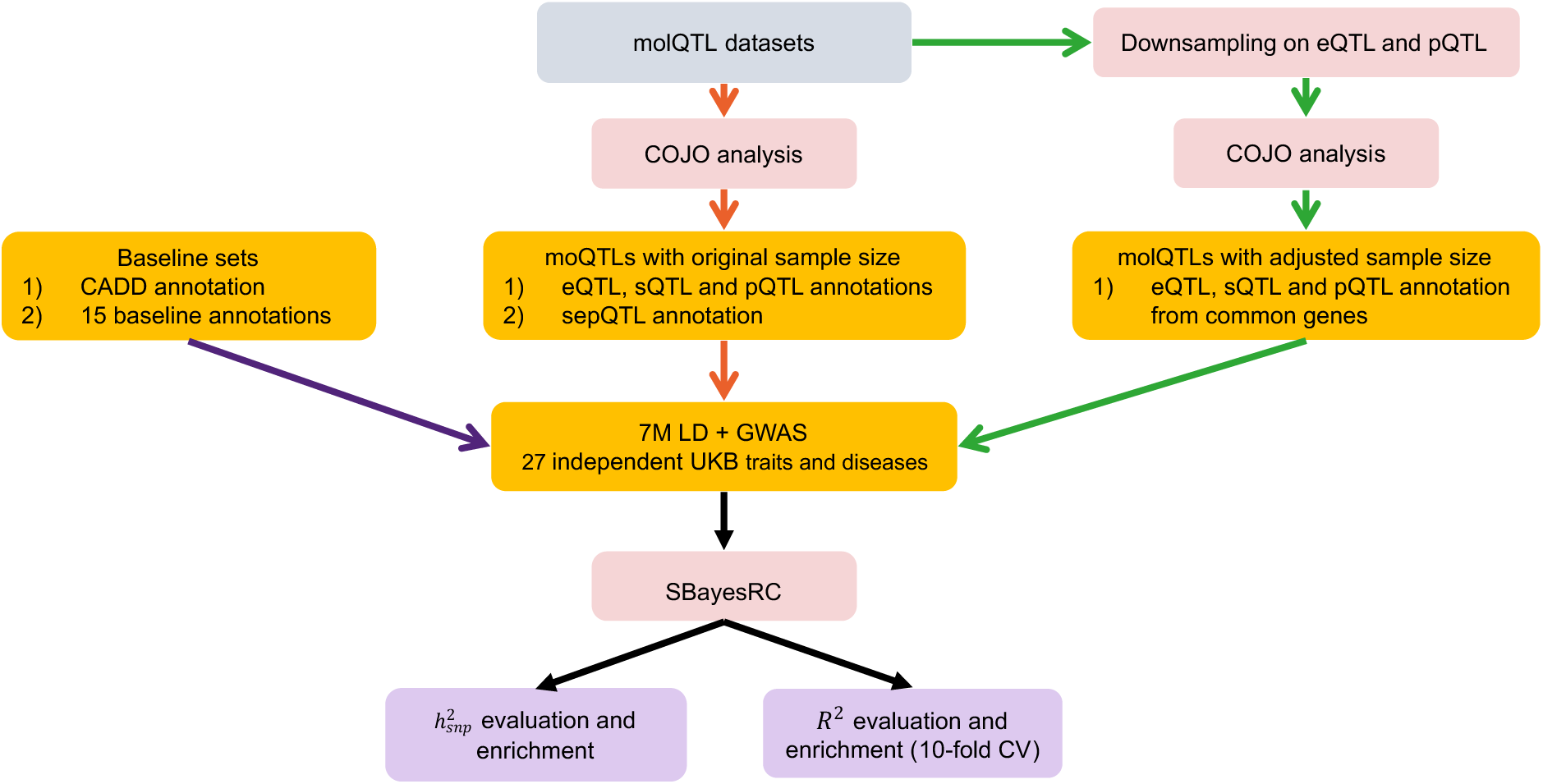
Study schema. Overview of the analytical workflow. We first generated eQTL, sQTL, and pQTL annotations, as well as their union, based on COJO analysis. For benchmark, we created a binary CADD annotation using the functional CADD meta-score and incorporated 15 additional functional annotations from the LDSC baseline annotation set. To account for differences in sample size and gene coverage, we applied a down-sampling strategy to the molQTL data in common genes across datasets, followed by COJO analysis to construct adjusted molQTL annotations. Finally, we used SBayesRC with multiple sets of molQTL annotations to evaluate their contributions to SNP-based heritability (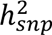) and prediction accuracy (𝑅^$^), as well as their enrichment across 27 GWAS traits and diseases.

**Table 1.** molQTL dataset used in the analysis.

| Date type | Resource | No. molecules | Mean No. cis-SNPs/molecule | Max sample size |
| --- | --- | --- | --- | --- |
| eQTL | eQTLGen consortium | 19,183 | 4,692 | 31,684 |
| sQTL | GTEx project | 64,999 | 4,861 | 670 |
| pQTL | UKB-PPP project | 2,856 | 5,193 | 34,090 |

## Results

### Contribution of the union of sQTL, eQTL, and pQTL (sepQTL) to SNP-based heritability

To explore the impact of sepQTL on SNP-based heritability estimation and enrichment, we applied SBayesRC, where the probability that a SNP affects a trait and the effect size of the SNP are estimated based on three sets of annotations: 1) the union of sQTL, eQTL, and pQTL (sepQTL) annotation, 2) the binarised CADD annotation (comprising SNPs with top CADD scores, matched in number to those in the sepQTL annotation), and 3) a set of 15 core functional annotations (Supplementary Table 4). These three sets of annotations (17 annotations in total) were each fitted in a separate SBayesRC analysis. Across the whole genome, the proportion of SNPs within each of the 17 annotations varied from 0.3% to 45.2%, with a median value of 1.8%, similar to that of the sepQTL annotations (1.2%). Despite the variation in overlap between annotations (Fig. 2a), the sepQTL are largely independent of most of these functional annotations, especially of the binarised CADD used for benchmarking (Fig. 2a).

**Figure 2.**
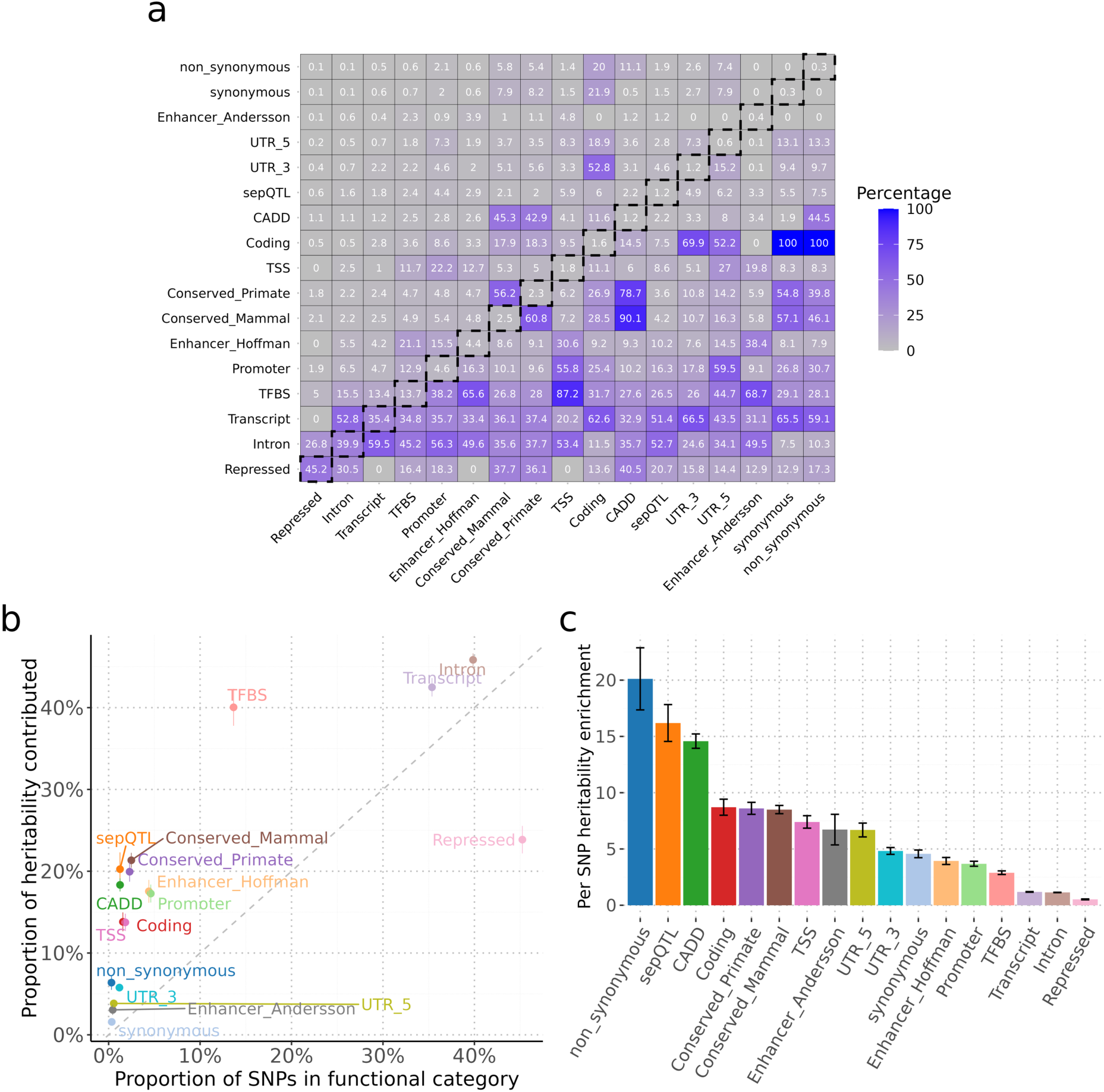
SNP-based heritability estimation and enrichment in sepQTL, CADD and other 15 functional categories across 27 independent UKB traits. **a)** Annotation overlap matrix based on Jaccard-like directional similarity metrics among sepQTL, CADD, and 15 additional functional annotations across 7.4 million genome-wide SNPs. The framed diagonal entries represent the proportion of SNPs annotated in each functional category. Annotation names are ordered according to the proportion of SNPs they annotate. The upper triangle (above the diagonal) shows 𝐗 ∩ 𝐘⁄𝐗— the proportion of overlap normalised by the column feature (𝐗), and the lower triangle (below the diagonal) shows 𝐗 ∩ 𝐘⁄𝐘— normalised by the row feature (𝐘). All values are shown as percentages. b) The relationship between the proportion of SNPs in functional categories and the proportion of heritability contributed by those functional categories. c) The distribution of per-SNP heritability enrichment across 17 annotations. Error bars represent standard errors across 27 traits.

Our results showed that sepQTL, although covering only 1.2% of the SNPs, accounted for 20.3% of the SNP-based heritability (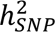), which is defined as the proprotion of phenotypic variance explained by all SNPs (Fig. 2b), leading to a 16.2× enrichment in the per-SNP 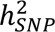 (Fig. 2c). These values were slightly higher than those of the binary CADD annotation, which had the same annotation size as sepQTL (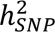 explained by SNPs with high CADD = 18.3%, per-SNP 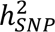 enrichment = 14.6×), and surpassed most other functional categories.

For comparison, coding regions had a slightly higher proportion of SNPs (1.6%) than sepQTL but explained a lower proportion of 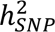 (13.8%) and therefore a lower per-SNP heritability enrichment (8.7×). The non-synonymous variants category was the only annotation with a greater per-SNP heritability enrichment than sepQTL (20.1× vs. 16.2×, with 6.4% of 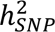 explained by 0.3% SNPs), which is not surprising given their direct functional relevance to amino acid coding in proteins. Conserved regions also showed an important enrichment (∼8×), explaining 20% of 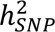 by 2.3% of SNPs, with consistent results across primate and mammalian conservation. Broad annotations like transcription factor binding sites (TFBS), transcript, and intron each explained over 40% of 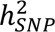 but had low per-SNP heritability enrichment (<3×), indicating that these annotations include many SNPs with small effects. As expected, although repressed regions covered 45.2% of the SNPs, they only accounted for 23.9% of 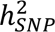, resulting in a depletion in per-SNP heritability enrichment (0.5×).

Overall, the strong enrichments derived from non-synonymous variants and sepQTL, followed by coding regions and transcription start sites (TSS), highlight the crucial roles of gene-regulatory and protein-altering variants in complex trait variation, comparable to the impact of predicted deleterious variants.

### Contribution of sepQTL to polygenic prediction

We further investigated the role of sepQTL in polygenic prediction accuracy and per-SNP predictability enrichment, in comparison with the CADD annotation and a set of 15 core functional genomic annotations (Fig. 3 and Supplementary Table 4). sepQTL contributed 34.2% of total prediction accuracy (measured as 𝑅^2^ based on the sepQTL as a fraction of that based on all SNPs), with a high per-SNP enrichment of 27.4× (Fig. 3a), comparable to that of the CADD annotation (27.7×) and exceeding all other annotations (0.7× ∼ 18.1×), except for non-synonymous variants (42.4×).

**Figure 3.**
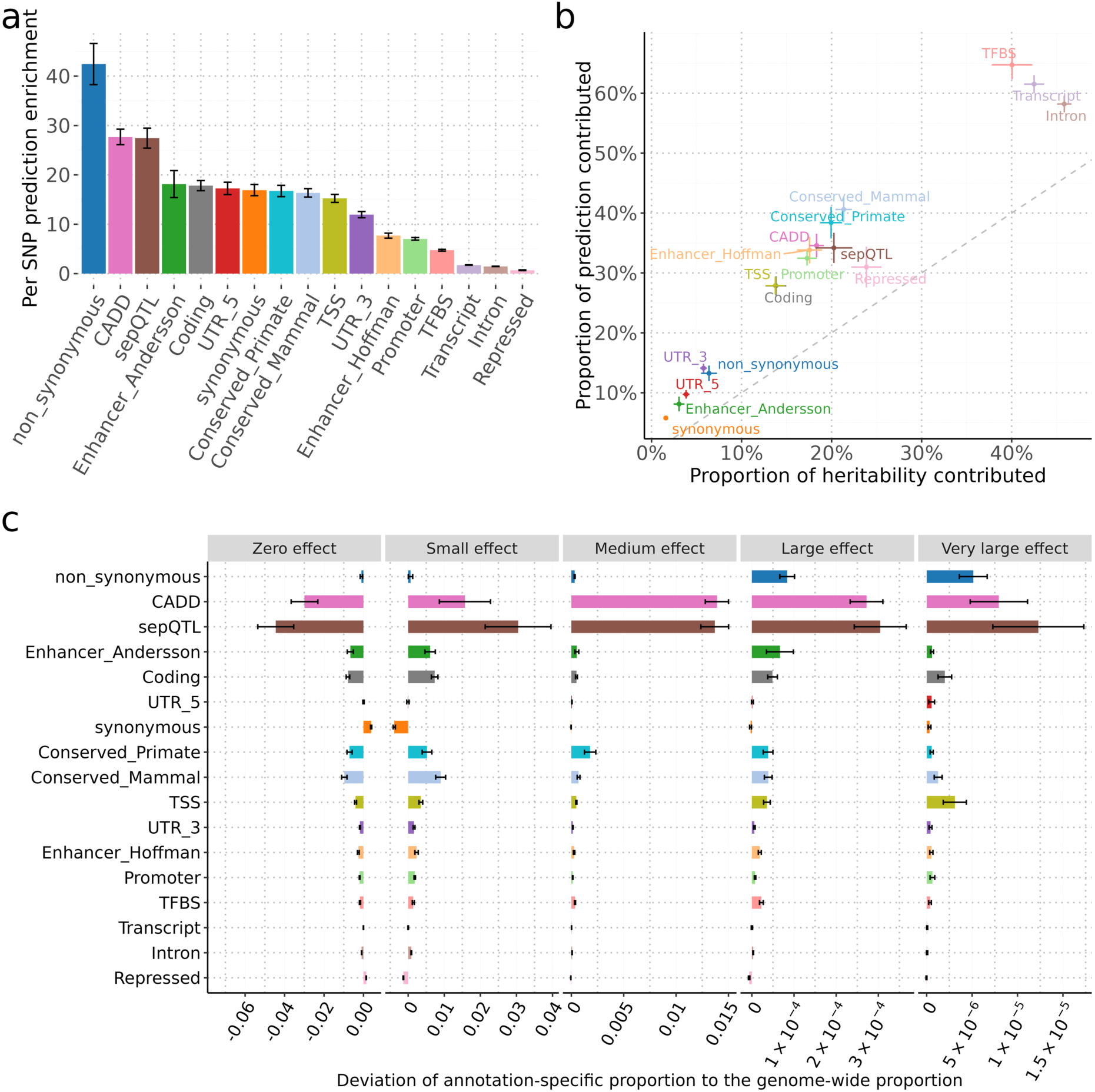
Proportions of polygenic prediction accuracy and enrichment in sepQTL, CADD, and other functional categories across 27 independent UKB traits. a) The distribution of per-SNP prediction enrichment across 17 annotations. Colours and shapes represent functional annotations, and error bars indicate standard errors (s.e.). b) Relationship between the proportion of 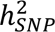 and the proportion of prediction contributed by different functional categories. The grey dash diagonal line is y=x. c) Distribution of effect sizes corresponding to the five mixture components in the Bayesian model. For each functional category, the distribution of effect sizes is illustrated by the deviations in the proportion of SNP effects assigned to each of the five mixture distributions to the overall proportion of genome-wide SNPs across 17 functional categories spanning 27 traits. SNP effect sizes are modelled as a mixture of five components with scaling factors 𝛾 = [0, 0.001, 0.01, 0.1, 1]% of 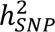, corresponding to zero, small, medium, large, and very large effects, respectively. Each component scales the total 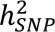 to represent different effect size magnitudes. Error bars represent standard errors across 27 traits.

Although conserved regions contributed a larger proportion of prediction 𝑅^2^ (40.6% in mammalian conservation, 38.4% in primate conservation), their per-SNP predictability enrichment was lower (∼16× each). This suggests that while conserved regions contain many functionally important variants, their individual predictive power is lower than that of sepQTL. UTR regions also exhibited significant contributions to prediction (14.1% in 3’-UTR, 9.7% in 5’-UTR), but their per-SNP enrichment (11.9× in 3’-UTR, 17.3× in 5’-UTR) remained lower than that of sepQTL and CADD annotations. Large annotation categories, such as TFBS, Transcript, and Intron, contributed substantially to total prediction 𝑅^2^ (up to 60%) but showed modest per-SNP predictability enrichments (ranging from 1.5x to 4.7×), indicating weaker effect sizes. These results highlight the high predictive value of sepQTL, placing them among the most enriched functional categories alongside CADD and non-synonymous variants. Unlike broad regulatory annotations, which include many small-effect SNPs, sepQTL are enriched with large-effect regulatory variants critical for genomic prediction.

Interestingly, the prediction 𝑅^2^ contributions and enrichments across all annotations exceeded their corresponding heritability estimates (Fig. 3b). For instance, the non-synonymous annotation accounted for 13.2% of total prediction 𝑅^2^ and exhibited an enrichment of 42.4×, more than twice its corresponding heritability proportion. This discrepancy can be attributed to the differential shrinkage of SNP effects in the SBayesRC model, where small effects are shrunk more towards zero while large effects are shrunk less. As shown in functional architecture estimated from SBayesRC (Fig. 3c and Supplementary Table 5), sepQTL showed the lowest proportion of SNPs with zero effect comparing to the other annotation categories (about 5% lower than the genome-wide estimate), and the highest proportions of SNPs in non-zero effect size categories, especially in the very large effect category (explaining 1% of the 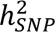), highlighting their strong relevance to trait genetic variation. While high-CADD and non-synonymous SNPs also showed enrichment for large effects, the enrichment was strongest for sepQTL. In contrast, evolutionary conserved regions and enhancer regions tended to be more enriched in the small-effect category. These findings support the effectiveness of molQTL-based annotations in capturing variants with large effects, highlighting their utility in polygenic prediction.

### Relative contributions of eQTL, sQTL, and pQTL to complex trait variation

We have assessed the contribution of eQTL, sQTL, and pQTL as a whole (sepQTL) to 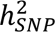 and polygenic prediction. We next compare their relative contributions by creating an annotation for each of these molQTL and considering them jointly in the SBayesRC model. Although these molQTL each contained a very small fraction of the genome, they have all made disproportionally large contributions to 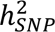 and prediction accuracy (Fig. 4a). Specifically, approximately 1.0% of SNPs were eQTL, explaining 13.9% of 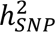 and contributing to 25.5% of prediction accuracy, on average across traits. For sQTL (pQTL), we observed that 0.1% (0.2%) SNPs explained 2.8% (4.0%) of 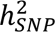 and contributed to 7.0% (8.0%) of prediction 𝑅^2^.

**Figure 4.**
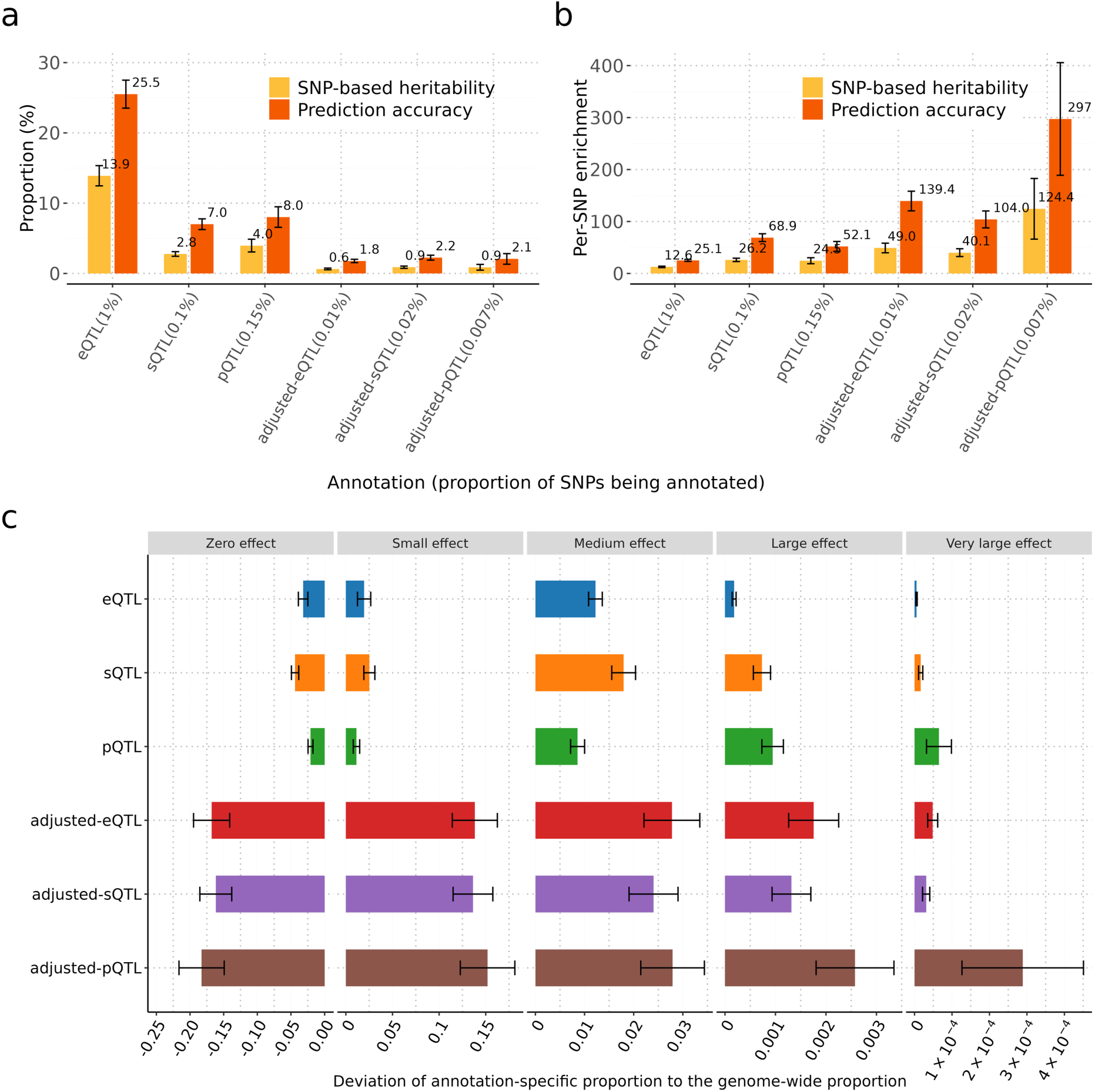
Comparison of contributions of eQTL, sQTL, and pQTL on SNP-based heritability and polygenic prediction. a) Relationship between the proportion of 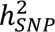 and the proportion of prediction accuracy contributed by eQTL, sQTL, and pQTL annotations with original and adjusted sample sizes. Yellow bars indicate the proportion of 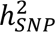 explained by SNPs within each functional annotation category. Orange bars represent the corresponding prediction accuracy. Error bars denote standard errors across 27 traits. Functional categories are labelled along the x-axis with the percentage of SNPs covered in each. b) Per-SNP heritability and prediction shown by the proportion of SNPs (x-axis) across molQTL-related annotation sets. c) Distribution of effect sizes belonging to each of the five mixture distribution components. For each functional category, the distribution of effect sizes is illustrated by the deviations in the proportion of SNP effects assigned to each of the five mixture distributions from the overall proportion of genome-wide SNPs across 17 functional categories spanning 27 traits. SNP effect sizes are modelled as a mixture of five components with scaling factors 𝛾 = [0, 0.001, 0.01, 0.1, 1], corresponding to zero, small, medium, large, and very large effects, respectively. Each component scales the total 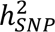 to represent different effect size magnitudes.

These comparisons, however, are confounded by differences in sample size and genome coverage across molQTL datasets. For example, the eQTL data we used had the largest sample size but was limited to protein-coding genes, whereas the sQTL data had the smallest sample size but covered a broader set of genes. The pQTL data were restricted to proteins measured in the Olink platform in the UKB. These disparities in sample size and gene coverage would influence statistical power and introduce ascertainment bias in molQTL comparisons.

To address this, we selected 1,457 genes in common across datasets and applied a down-sampling approach to match the sample sizes of eQTL and pQTL data to that of the sQTL data (Methods). Simulation analyses confirmed that our down-sampling method produces eQTL effect size and standard error estimates that are consistent with those directly obtained from the target sample size (Supplementary Figure 1). As sample size decreased, eQTL maintained similar effect sizes but exhibited larger standard errors. After adjustment, the proportion of SNPs with eQTL or pQTL annotations decreased markedly, from 1.0% to 0.013% for eQTL and 0.15% to 0.007% for pQTL (Supplementary Table 6), and the number of independent QTL per gene declined from 4 to fewer than 1 for both eQTL and pQTL (Supplementary Figure 2).

After down-sampling and gene matching, we observed comparable contributions of eQTL, pQTL, and sQTL to both 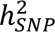 and prediction 𝑅^2^ (Fig. 4a and 4b). Interestingly, adjusted sQTL and pQTL tended to explain a slightly larger proportion of 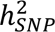 and prediction 𝑅^2^ than adjusted eQTL, consistent with recent findings emphasising their regulatory importance^16,17^. The ranking of prediction 𝑅^2^ contributions among different molQTL annotations closely followed that of 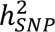, although the corresponding prediction 𝑅^2^ values were notably higher. This difference likely reflects the fact that most causal variants were enriched in groups with medium to large effect sizes (Fig. 4c and Supplementary Table 5), whereas variants with small effects were regressed more in the prediction.

### Variation of eQTL, sQTL, and pQTL contributions across complex traits

We examined the influence of different molQTL types on 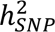 estimation and prediction 𝑅^2^ across 27 complex traits using SBayesRC with the original molQTL sample sizes (Fig. 5 and Supplementary Table 7). Our analysis revealed substantial variation in 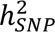 proportion and enrichment estimates across traits (Fig. 5a,b). On average, sepQTL explained 20.3% of 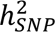, with the highest contributions for blood-related traits (29.9%), followed by disease traits (22.9%), but lower for behaviour and cognitive traits, implying tissue specificity of molQTL effects.

**Figure 5.**
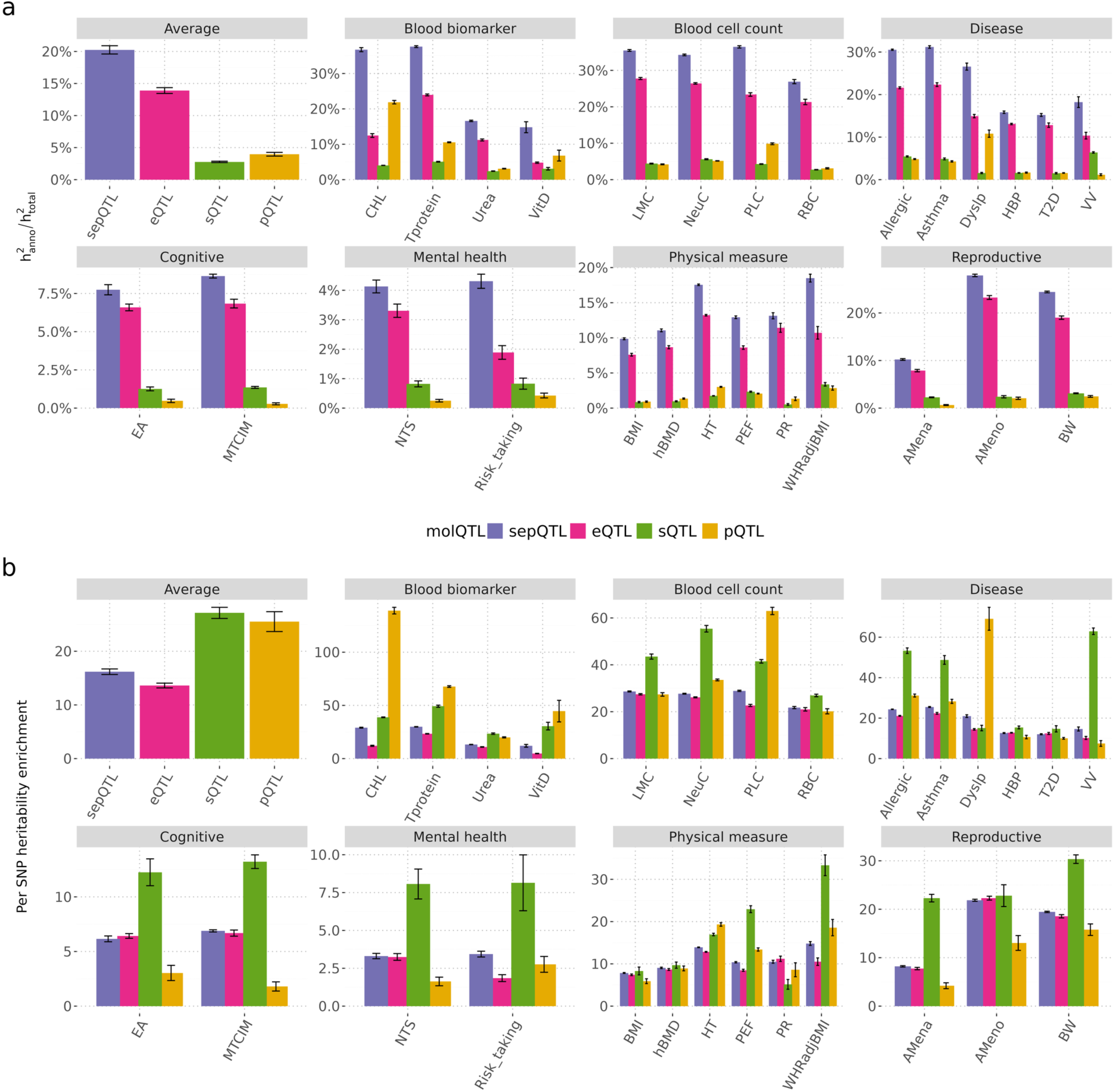
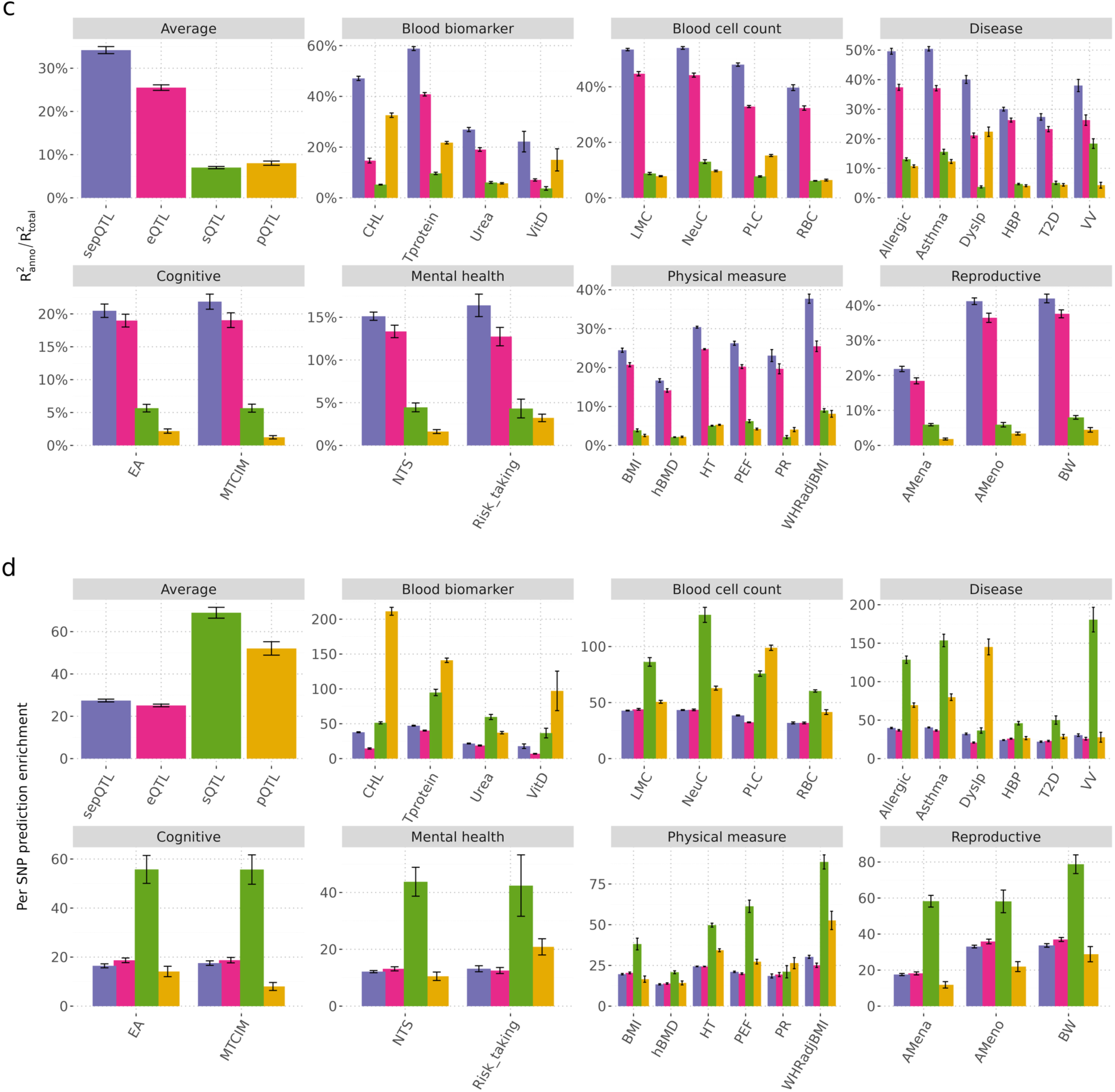
Variation of molQTL contributions to SNP-based heritability and prediction accuracy across complex traits, with the original sample sizes for molQTL datasets. a) 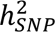 proportion and b) per-SNP heritability enrichment contributed by different molQTL; c) prediction accuracy proportion and d) per-SNP prediction enrichment contributed by different molQTL. 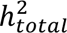 refers to 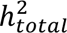 using SNPs in the whole genome. 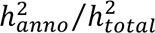 refers to the proportion of heritability contributed by molQTL annotations. The average category refers to corresponding average values across traits in different models. 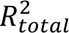 refers to the total prediction 𝑅^2^ using SNPs in the whole genome. 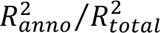 refers to the proportion of prediction 𝑅^2^ contributed by molQTL annotations. The average category refers to corresponding average values across traits in different models. Different colours represent distinct molQTL annotations. Error bars indicate standard errors across 27 traits.

Among individual molQTL types, eQTL contributed the most to 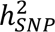 on average (13.9%), followed by pQTL (4.0%) and sQTL (2.8%). However, per-SNP heritability enrichment showed a distinct pattern: sQTL exhibited the highest enrichment (27.2×), followed by pQTL (25.5×) and eQTL (13.6×). The impact of molQTL on 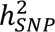 and enrichment was highly trait-specific. For example, eQTL explained 27.8% of 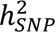 for lymphocyte count (LMC), whereas pQTL had the highest per-SNP heritability enrichment for cholesterol (CHL) at 139.1×. For total protein (TProtein), eQTL contributed 23.9% of 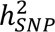 with 23.3× enrichment, while sQTL explained only 5.1% of 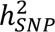 but showed a much stronger enrichment of 49.2×.

We next evaluated the contributions of molQTL to prediction *R^2^* across traits (Fig. 5c,d). Consistent with the 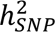 estimates, sepQTL explained the highest overall *R^2^* proportion (34.2% on average), with particularly strong contributions for blood-related traits (43.8%) and diseases (39.3%). While eQTL accounted for the largest overall prediction *R^2^* (25.5%), their per-SNP *R^2^* enrichment (25.1×) was lower than that of sQTL (68.9×) and pQTL (52.1×). Trait-specific enrichment patterns were again evident, for example, eQTL contributed most to prediction *R^2^*for LMC (44.7%, 44.0×), sQTL for varicose veins (VV; 18.4%, 181.0×), and pQTL for CHL (32.6%, 211.0×). Notably, blood-related and disease traits showed particularly high per-SNP predictability enrichment, aligning with their strong signals in heritability enrichment. On average across molQTL types, blood cell counts had a per-SNP predictability enrichment of 63.1×, while blood biomarkers reached 67.5×, underscoring the key role of context-specific regulatory variation in driving complex trait prediction.

## Discussion

In this study, we systematically evaluated the contributions of three types of molQTL in blood (eQTL, sQTL, and pQTL) to 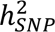 and prediction *R^2^* using SBayesRC. These molQTL types capture distinct layers of cis-regulatory genetic control at transcriptional and translational levels, providing functional insights into the genetic architecture of complex traits. We found that despite covering only 1.2% of SNPs, sepQTL (the union of the three molQTL types) explained 34.2% of total prediction *R^2^* with a 27.4× per-SNP enrichment, and 20.3% of 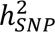 with a 16.2× enrichment, comparable to the contribution of predicted deleterious variants and outweighing 15 other functional annotations, including coding, enhancer, and conserved regions (Fig. 2 and 3). The central role of sepQTL was further supported by the annotation-stratified effect size architecture estimated by SBayesRC (Fig. 2c and 3c), where sepQTL had the highest proportion of nonzero-effect SNPs, particularly those with large effects (explaining ∼1% of 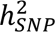). To enable fair comparisons between molQTL types, we developed a down-sampling method based on summary statistics, and found that given an equal sample size in the same gene set, sQTL and pQTL contributed slightly more to 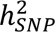 and prediction *R^2^* than eQTL (Fig. 4a). Notably, pQTL showed the strongest per-SNP heritability and predictability enrichment (Fig. 4b), likely driven by a greater proportion of SNPs with large effect sizes (Fig. 4c). These results highlight the important roles of post-transcriptional and translational regulation in shaping complex trait variation. Finally, we showed that the impact of these molQTL varied substantially across traits (contributing up to 13.9% of 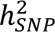 and 25.5% of prediction *R^2^*), with the largest contributions observed for blood-related traits (on average, eQTL contributed 18.9% 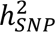 and 29.5% prediction *R^2^*) and diseases (eQTL contributed 15.9% 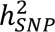 and 28.6% prediction *R^2^*), and the smallest for cognitive and mental health traits (Fig. 5). These results suggest that regulatory QTL for gene products may act in a tissue-specific manner, emphasizing the value of context-dependent molQTL annotations in trait heritability and polygenic prediction studies.

Several studies have assessed the role of eQTL in the genetic architecture of complex traits, but their contribution to trait heritability was found to be surprisingly limited. While 88% of genes expressed in blood have cis-eQTL^2^, Yao et al^15^ estimated that only ∼11% of trait heritability was mediated by eQTL across 42 complex traits. Hormozdiari et al.^19^ used LDSC and reported a ∼5.8× enrichment for eQTL annotation, which was less than that for exons and conserved regions^9,29^. In comparison, we estimated a similar proportion of 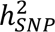 (13.9%) explained by eQTL but a greater fold enrichment (13.6×) (Fig. 4). The differences may be attributed to both the eQTL selection and the statistical methodology. Hormozdiari et al. constructed eQTL annotations from fine-mapping causal posterior probability (CPP), retaining the top 1% of SNPs, whereas we selected COJO-significant SNPs (also about 1% of SNPs), which may better capture common variation, as CPP can spread on SNPs in high LD with the causal variant and therefore rare variants would tend to be favoured. Both Yao et al and Hormozdiari et al relied on LDSC framework, which assumes an infinitesimal model, whereas we used SBayesRC, which models sparse genetic architectures and allows the distribution of effect sizes to vary across annotation categories. Despite a stronger role of eQTL was shown in our study, all these studies converge on an important insight, i.e., transcriptional regulation explains only part of the genetic basis of complex traits.

By incorporating additional regulatory layers, i.e., sQTL and pQTL, we showed that sepQTL can collectively account for a substantial proportion of 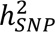 (20.3%). This observation is consistent with Xiang et al.^21^, who quantified the impact of LD-pruned eQTL and sQTL on the genetic architecture of 37 complex traits in cattle using BayesRC, a similar model to SBayesRC but requires individual-level data. They found that cis QTL affecting both expression and splicing explained 12.7% of 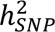 in single-tissue analyses, rising to 45.3% in multi-tissue analyses and 69.2% when including trans effects, highlighting the potential of multi-layer and multi-tissue molQTL integration to capture regulatory effects that are underrepresented in conventional annotation-based heritability models.

Our down-sampling method of molecular datasets enabled direct comparisons between molQTL types. Our results showed that the proportion of 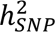 explained increased with the size of the molecular dataset. Therefore, it is likely that with larger datasets, sepQTL would explain more of 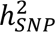. Xiang et al used multiple tissues as well as cis and trans eQTL, and this may also explain the larger fraction of 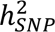 that they explained by seQTL. As molQTL datasets continue to increase in sample size and cell type diversity, we anticipate future benchmarking of molQTL types at cellular resolution.

Another contribution of our work is in polygenic prediction. Polygenic prediction models that incorporate functional annotations have shown improved prediction accuracy both within and between ancestries^30–34^. We showed that sepQTL contributed substantially to total prediction *R^2^*, with greater per-SNP enrichment for predictability than for heritability. This is likely because the overrepresentation of large-effect SNPs in sepQTL, which are regressed less toward zero compared to the small-effect SNPs. In this study, we focused on evaluating the contribution of sepQTL to prediction accuracy. However, the extent to which incorporating cell-type-specific or context-dependent QTL can improve the overall prediction performance remains an important area for future research.

This study has several limitations. First, we selected only significant molQTL filtered by COJO and created binary molQTL annotations based solely on their presence or absence. This approach may result in an overrepresentation of large-effect molQTL while neglecting weaker yet potentially meaningful genetic signals. Future research should explore effect size-weighted annotations, where molQTL effect sizes are integrated as continuous weights into Bayesian models like SBayesRC to improve the accuracy of functional SNP impact predictions. Second, our analysis primarily focused on cis-QTL without considering trans-effects. This limitation may overlook important regulatory interactions occurring at distal genomic regions^2,21^, which could play a crucial role in mediating genetic influences on complex traits. Third, we investigated the non-infinitesimal genetic architectures of 27 complex traits and observed that SNPs with large and very large effect sizes were particularly enriched in pQTL-associated regions. This suggests that pQTL may tag highly functional genetic elements that influence phenotypic variation. Further studies are required to elucidate the biological mechanisms underlying the distinct enrichment patterns observed across different molQTL annotations. Fourth, our findings highlight the necessity of analysing molQTL data from trait-relevant tissues, particularly in multi-omics studies that seek to disentangle the molecular mechanisms of genetic risk. Accounting for tissue specificity may enhance the biological relevance of molQTL-based analyses and improve the interpretation of genetic regulation across different molecular layers.

In conclusion, our findings demonstrate the substantial contributions of expression, splicing, and protein QTL to genetic variation and polygenic prediction across complex traits. As genomic datasets continue to expand, integrating diverse molQTL annotations across tissues and molecular layers will be critical for enhancing genetic risk prediction, improving fine-mapping strategies, and uncovering novel biological pathways underlying complex traits.

## Methods

### Ethics approval

The University of Queensland Human Research Ethics Committee B (2011001173) provides approval for analysis of human genetic data used in this study on the high-performance cluster of the University of Queensland.

### GWAS data

Following previous studies^30^, we selected 27 independent traits from the UK Biobank dataset, restricted to 341,809 unrelated individuals of European descent (Supplementary Table 1). For continuous phenotypes, extreme values beyond ±7 standard deviations from the mean were excluded, followed by a rank-based inverse normal transformation, applied separately within European ancestry and sex groups. Genotype quality control involved removing multi-allelic variants, as well as SNPs with imputation information scores < 0.3, hard-call genotype probability < 0.9, minor allele frequency (MAF) < 0.01, Hardy–Weinberg equilibrium p-values < 10⁻⁶, or missingness rates > 1%. After filtering, approximately 7 million (7,356,518) high-quality SNPs were retained for downstream analysis. To define the cis-region for a gene, we mapped SNPs to a flanking window of 1MB on both sides of the gene midpoint position based on GENCODE (v40, https://www.gencodegenes.org/human/release_40.html) under the hg19 genome build (50KB was used as the flanking window size in simulations).

### molQTL datasets

This study incorporated three types of cis-molQTL datasets, defining the cis region for each gene as a 2 megabase (2 Mb) window centered at the gene’s midpoint (±1 Mb) (Table 1). eQTL data were sourced from the eQTLGen consortium^2^, which performed a blood-based meta-analysis across approximately 6 million cis-SNPs and 19,183 genes, based on an average sample size of 20,519 individuals. pQTL data were obtained from The Pharma Proteomics Project^7^, which profiled nearly 4 million cis-SNPs in relation to 2,856 plasma proteins, using a cohort with an average sample size of 33,394. sQTL data came from the GTEx v8 resource^3^ (n = 670 whole blood samples), comprising ∼6 million cis-SNPs and 65,127 genes (Table 1 and Supplementary Table 2). The differences in gene counts across molQTL datasets reflect both biological distinctions among QTL types and technical heterogeneity in discovery platforms (Supplementary Table 3). For instance, the GTEx sQTL dataset includes 65,127 entries corresponding to splice junctions rather than genes, accounting for its large size. In contrast, the eQTLGen consortium reported cis-eQTL for approximately 19,183 protein-coding genes, while the pQTL dataset derived from the UK Biobank-PPP Olink platform includes 2,856 proteins.

To enable fair comparisons across molQTL types, we identified a set of 1,457 genes that are shared across all three molQTL datasets based on Ensembl gene IDs. We then applied a down-sampling strategy to match the sample sizes of eQTL and pQTL datasets to that of the sQTL dataset, reducing potential power-related biases. After performing COJO analysis on the down-sampled data, we retained 973 genes with a total of 2,966 molQTL. These harmonised annotations are referred to as adjusted-molQTL, representing a comparable and balanced dataset for downstream analyses.

### COJO analysis

To evaluate the contribution of molQTL to 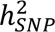 and prediction accuracy, we constructed several molQTL binary annotations. We applied conditional and joint association analysis (COJO)^35^ to the summary statistics of each type of molQTL (eQTL, sQTL, and pQTL) to identify conditionally independent molQTL signals with a p-value < 5 × 10^%&^. SNPs identified as significant molQTL by COJO (referred to as molQTL hereafter for brevity) were assigned a value of one, while all other SNPs were assigned zero, thereby constructing separate molQTL annotations. In addition, a binary annotation was generated for SNPs that were at least one of the three types of molQTL, referred to as the sepQTL annotations.

To account for sample size differences among molQTL types, we applied a summary-data-based down-sampling approach to the conditionally independent eQTL and pQTL to ensure their sample sizes matched that of sQTL. Following this, we conducted COJO analysis for 1457 common genes and identified molQTL for 973 genes in total across the three molQTL datasets to construct adjusted-molQTL annotations.

## Functional annotations for benchmarking

To assess the relative importance of the sepQTL annotations compared to evolutionary genomic annotations, which have been found to be highly enriched in per-SNP heritability for complex traits^30^, we defined a CADD binary annotation by selecting SNPs in the top 1.25% of CADD scores, matching the proportion of sepQTL. SNPs above this threshold were assigned a value of one, while all others were assigned zero.

For comparison, a set of 15 blood-related functional annotations was derived from the LDSC BaselineLD model, including coding, Intron, Promoter, 3’-UTR (UTR_3), 5’-UTR (UTR_5), Transcript (Transcriptional Regulatory Elements identified in studies led by Michael M. Hoffman), Enhancer_Andersson, Enhancer_Hoffman, Repressed, TFBS (Transcription Factor Binding Sites identified by the Encyclopedia of DNA Elements Project), TSS (Transcription Start Sites identified in studies led by Michael M. Hoffman), synonymous, non_synonymous, Conserved_Mammal (a conservation score derived from 46-way multiple sequence alignment across vertebrate species, with a focus on mammals), Conserved_Primate (a conservation score calculated using the phastCons algorithm on a 46-way multiple sequence alignment, with a focus on primate species).

### SBayesRC analysis

SBayesRC is a Bayesian framework developed for estimating 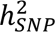 and improving polygenic prediction by integrating GWAS summary statistics and functional annotations^30^. It assumes that SNP effect sizes follow a five-component mixture of normal distributions, each reflecting varying levels of genetic contribution, ranging from no effect to very large effects (e.g., 0, 0.001, 0.01, 0.1, and 1). The proportion assigned to each component is inferred from the data and modulated by relevant genomic features, such as regulatory elements or conservation status. The SBayesRC model used here was

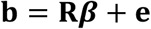

Where 𝐛 denotes GWAS marginal effect vector, the LD matrix denotes defined as 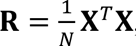, 𝑁 denotes the GWAS N sample size, 𝜷 denotes SNP joint effect vector, and 𝐞 denotes the residual vector.

For the joint effect size of SNP 𝑗,

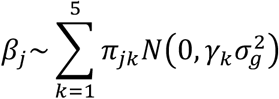

Where 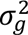 reflects the overall genetic variance attributable to SNPs, as estimated from the dataset. The vector 𝛾 = [0, 0.001, 0.01, 0.1, 1]^(^% specifies the scaling factors corresponding to the five components in the mixture model. This mixture comprises a point mass at zero alongside four Gaussian components, with each SNP assumed a priori to contribute between 0.001% and 1% to the total genetic variance. The parameter 𝜋*_jk_* denotes the probability that a given SNP belongs to the 𝑘-th distribution.

When considering the influence of annotations, for each SNP, we further model 𝜋*_jk_* as

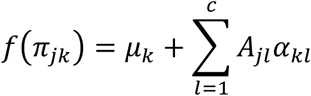

where 𝑓(·) serves as a link function, transforming the probability parameter 𝜋*_jk_* onto the real line, 𝜇*_k_* denotes the baseline log-probability for 𝑘-th distribution, *A_jl_* denotes the value of annotation 𝑙 on SNP 𝑗, and *α_kl_*. denotes the effect of annotation 𝑙 on the membership probability to the 𝑘 -th distribution. In real analysis, block-diagonal LD matrices were employed, each spanning at least 4 cM, ultimately yielding 591 combined regions among individuals of European descent.

### SNP-based heritability partitioning and enrichment estimation

The total SNP heritability for a trait is defined as the variance of the trait explained by the genetic variance (assuming unit phenotypic variance):

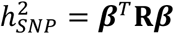

And the annotation-specific heritability of a binary annotation 𝑐 is defined by the variance of the trait explained by the SNPs within the annotation:

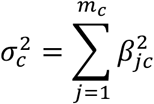

Where 𝑚_/_ is the number of SNPs within the annotation.

We ran SBayesRC for 27 independent UKB complex traits and diseases to evaluate 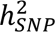, incorporating five sets of molQTL-related annotations: 1) a set of three separate moQTL annotations, 2) a sepQTL annotation set alone, 3) a set of three separate adjusted-molQTL annotations in common genes, 4) a binary CADD annotation set alone, 5) a set of 15 core functional annotations. To account for differences in SNP density across annotations, we assessed per-SNP heritability enrichment. In line with the framework of stratified LD score regression, heritability enrichment for a given annotation is defined as the proportion of 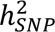 explained by that annotation, divided by the proportion of SNPs it encompasses.

### Prediction accuracy partitioning and enrichment estimation

To evaluate predictive performance, a 10-fold cross-validation framework was applied to the UK Biobank dataset, involving 341,809 unrelated individuals of European ancestry. For each fold, 90% of the samples were used to train the model, while the remaining 10% served as the validation set. GWAS summary statistics were computed using PLINK2 software^36^, adjusting for key covariates, including sex, age, and the top 10 genetic principal components. Trait-specific regression models were employed, with linear regression applied to continuous outcomes and logistic regression used for binary traits. Prediction accuracy enrichment is calculated as the proportion of total prediction accuracy attributed to the annotation, normalised by its SNP count proportion.

### Down-sampling molQTL summary statistics

There were large differences in the sample size among the three types of molQTL, with sQTL having the smallest (N_sQTL_=670)^18^. To address this issue, we developed a method to estimate the summary statistics of eQTL and pQTL with down-sampled sample sizes matched with that of the sQTL study. For a given gene (or protein), we defined 𝑏*_j_^(c)^* as the molQTL effect for SNP 𝑗 with the current sample size and 𝑏*_j_*^(*t*)^ as that with the target sample size. Under the assumption of normality and unit phenotypic variance, the joint distribution of 𝑏*_j_*^(*c*)^ and 𝑏*_j_*^(*t*)^can be approximated by a bivariate normal distribution. For each gene, given the marginal effect-size vector 𝐛^(*c*)^ and LD matrix 𝐑, the conditional distribution 𝐛^(*t*)^|𝐛^(*c*)^ is:

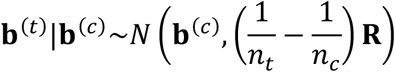

where 𝑛_/_ and 𝑛_3_are the current and target sample sizes for each molQTL within that gene, respectively, with 𝐛^(*c*)^used as a surrogate of the true effect sizes. Eigen-decomposition with a maximum cut-off of variance explained (𝜌 = 0.995) was applied to the LD matrix to generate a pseudo-inverse of 𝐑. The summary statistics under the target sample size were then estimated as

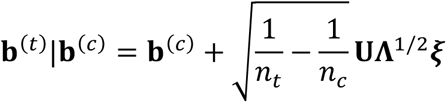

where 𝐔 and 𝚲 are matrices of eigenvectors and eigenvalues of the LD matrix 𝐑, and 𝜉*_j_*∼𝑁(0,1) is a standard normal random number for SNP 𝑗. The marginal standard error (**SE**) for 𝐛^(*t*)^and 𝐛^(*c*)^is 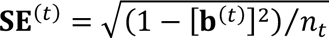 and 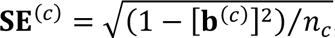, respectively. Equivalently, 𝐒𝐄^(*t*)^can be estimated from 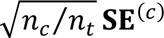.

### Simulations

To evaluate how effective the proposed summary-level down-sampling method is, we generated simulated gene expression and quantitative traits based on genotypic data sourced from the UK Biobank resource. The analysis was restricted to HapMap 3 SNPs on chromosome 22, comprising 12,159 SNPs, and included a randomly selected set of 50,000 unrelated participants of European ancestry.

We conducted a simulation study where SNPs influenced a complex trait through gene expression. Specifically, we randomly selected 10 genes as causal, each associated with approximately 10 causal SNPs drawn from a Poisson distribution (mean = 10). These SNPs were sampled within a 50-kilobase (kb) window centred around each gene’s midpoint. Their effect sizes were sampled from a standard normal distribution. Expression levels were simulated for 50,000 individuals under a linear additive model, incorporating random environmental noise. The heritability of expression was set to 50%, defined as the proportion of variance attributable to genetic components across genes. Phenotypes were then generated based on the simulated expression levels, again using an additive model with Gaussian noise, where gene expression explained 10% of the total trait variance. Throughout the simulation, centred and scaled genotypes were used, and both gene expression levels and trait phenotypes were centred and scaled. GWAS and eQTL marginal summary statistics were estimated using simple linear regression. The LD reference matrix was computed using internal genotype data.

To evaluate how sample size impacts the estimation of eQTL effect sizes and their standard errors, we performed down-sampling for each gene across three experimental settings: **a**) the full dataset with 50,000 individuals, **b**) a moderate-sized subset of 10,000, and **c**) a small-scale cohort of 1,000 samples.

## Data availability

No data were generated in the present study. The eQTL summary statistics are publicly available at https://www.eqtlgen.org/. The sQTL summary statistics are publicly available at https://yanglab.westlake.edu.cn/software/smr/#DataResource. The pQTL summary statistics are available at https://www.synapse.org/Synapse:syn51364943/. The UK Biobank data are from the UK Biobank resource at https://www.ukbiobank.ac.uk/enable-your-research/about-our-data/.

## Code availability

For plink2.0, see https://www.cog-genomics.org/plink/2.0/. For GCTA-COJO, see https://yanglab.westlake.edu.cn/software/gcta/#COJO. For SBayesRC, see https://cnsgenomics.com/software/gctb/#SBayesRCTutorial. The methods related to calculating gene LD reference and down-sampling strategy are implemented in the C++ software BayesOmics, which is freely available at https://github.com/ShouyeLiu/BayesOmics.

## Data Availability

All data produced in the present study are available upon reasonable request to the authors
All data produced in the present work are contained in the manuscript

https://www.eqtlgen.org/

https://yanglab.westlake.edu.cn/software/smr/#DataResource

https://www.synapse.org/Synapse:syn51364943/

https://www.ukbiobank.ac.uk/enable-your-research/about-our-data/

## Acknowledgements

We thank the Research Computing Centre (RCC) Infrastructure Team at the University of Queensland for their support in this research. This research was supported by the Australian National Health and Medical Research Council (1177268) and the Australian Research Council (FL180100072). This study makes use of data from the UKB (project ID 12505).

## Author contributions

J.Z. conceived and designed the study. S.L. performed simulations and real data analysis under the assistance and guidance of X.Q., Y.W, Z.Z., M.E.G., J.Y., P.M.V., and J.Z. S.L. developed the software with the assistance of J.Z. and Z.Z. J.Z. and S.L. wrote the manuscript with the participation of all authors.

## Competing interests

The authors declare no competing interests.

## Additional information

Supplementary information is available for this paper at the link.

## Notes

### Competing Interest Statement

The authors have declared no competing interest.

### Clinical Protocols

https://shouyeliu.github.io/softwares/content-softwares.html

## Reference

1. Lappalainen, T., Li, Y.I., Ramachandran, S. & Gusev, A. Genetic and molecular architecture of complex traits. Cell 187, 1059–1075 (2024).

2. Võsa, U. et al. Large-scale cis-and trans-eQTL analyses identify thousands of genetic loci and polygenic scores that regulate blood gene expression. Nature genetics 53, 1300–1310 (2021).

3. Consortium, G. The GTEx Consortium atlas of genetic regulatory effects across human tissues. Science 369, 1318–1330 (2020).

4. Sun, B.B. et al. Genomic atlas of the human plasma proteome. Nature 558, 73–79 (2018).

5. Abascal, F. et al. Expanded encyclopaedias of DNA elements in the human and mouse genomes. Nature 583, 699–710 (2020).

6. Kerimov, N. et al. A compendium of uniformly processed human gene expression and splicing quantitative trait loci. Nature genetics 53, 1290–1299 (2021).

7. Sun, B.B. et al. Plasma proteomic associations with genetics and health in the UK Biobank. Nature, 1–10 (2023).

8. Zhu, H. & Zhou, X. Transcriptome-wide association studies: a view from Mendelian randomization. Quantitative Biology, 1–15 (2020).

9. Gusev, A. et al. Integrative approaches for large-scale transcriptome-wide association studies. Nature genetics 48, 245–252 (2016).

10. Giambartolomei, C. et al. Bayesian test for colocalisation between pairs of genetic association studies using summary statistics. PLoS genetics 10, e1004383 (2014).

11. Burgess, S., Butterworth, A. & Thompson, S.G. Mendelian randomization analysis with multiple genetic variants using summarized data. Genetic epidemiology 37, 658–665 (2013).

12. Chun, S. et al. Limited statistical evidence for shared genetic effects of eQTLs and autoimmune-disease-associated loci in three major immune-cell types. Nature genetics 49, 600–605 (2017).

13. Connally, N.J. et al. The missing link between genetic association and regulatory function. Elife 11, e74970 (2022).

14. Mostafavi, H., Spence, J.P., Naqvi, S. & Pritchard, J.K. Systematic differences in discovery of genetic effects on gene expression and complex traits. Nature Genetics 55, 1866–1875 (2023).

15. Yao, D.W., O’Connor, L.J., Price, A.L. & Gusev, A. Quantifying genetic effects on disease mediated by assayed gene expression levels. Nature Genetics 52, 626–633 (2020).

16. Li, Y.I. et al. RNA splicing is a primary link between genetic variation and disease. Science 352, 600–604 (2016).

17. Qi, T. et al. Genetic control of RNA splicing and its distinct role in complex trait variation. Nature genetics, 1–9 (2022).

18. Wu, Y. et al. Joint analysis of GWAS and multi-omics QTL summary statistics reveals a large fraction of GWAS signals shared with molecular phenotypes. Cell Genomics (2023).

19. Hormozdiari, F. et al. Leveraging molecular quantitative trait loci to understand the genetic architecture of diseases and complex traits. Nature genetics 50, 1041–1047 (2018).

20. Finucane, H.K. et al. Partitioning heritability by functional annotation using genome-wide association summary statistics. Nature genetics 47, 1228–1235 (2015).

21. Xiang, R. et al. Gene expression and RNA splicing explain large proportions of the heritability for complex traits in cattle. Cell Genomics 3(2023).

22. Brown, A.A. et al. Genetic analysis of blood molecular phenotypes reveals common properties in the regulatory networks affecting complex traits. Nature Communications 14, 5062 (2023).

23. Kim-Hellmuth, S. et al. Genetic regulatory effects modified by immune activation contribute to autoimmune disease associations. Nature Communications 8, 266 (2017).

24. Goode, D.L. et al. Evolutionary constraint facilitates interpretation of genetic variation in resequenced human genomes. Genome research 20, 301–310 (2010).

25. Zhu, X., Ma, S. & Wong, W.H. Genetic effects of sequence-conserved enhancer-like elements on human complex traits. Genome Biology 25, 1 (2024).

26. Sullivan, P.F. et al. Leveraging base-pair mammalian constraint to understand genetic variation and human disease. Science 380, eabn2937 (2023).

27. Rentzsch, P., Witten, D., Cooper, G.M., Shendure, J. & Kircher, M. CADD: predicting the deleteriousness of variants throughout the human genome. Nucleic acids research 47, D886–D894 (2019).

28. Bycroft, C. et al. The UK Biobank resource with deep phenotyping and genomic data. Nature 562, 203–209 (2018).

29. Barbeira, A.N. et al. Exploiting the GTEx resources to decipher the mechanisms at GWAS loci. Genome biology 22, 1–24 (2021).

30. Zheng, Z. et al. Leveraging functional genomic annotations and genome coverage to improve polygenic prediction of complex traits within and between ancestries. Nature Genetics 56, 767–777 (2024).

31. Hu, Y. et al. Leveraging functional annotations in genetic risk prediction for human complex diseases. PLoS computational biology 13, e1005589 (2017).

32. Hu, Y. et al. Joint modeling of genetically correlated diseases and functional annotations increases accuracy of polygenic risk prediction. PLoS genetics 13, e1006836 (2017).

33. Márquez-Luna, C. et al. Incorporating functional priors improves polygenic prediction accuracy in UK Biobank and 23andMe data sets. Nature Communications 12, 1–11 (2021).

34. Zhuang, Y., Kim, N.Y., Fritsche, L.G., Mukherjee, B. & Lee, S. Incorporating functional annotation with bilevel continuous shrinkage for polygenic risk prediction. BMC Bioinformatics 25, 65 (2024).

35. Yang, J. et al. Conditional and joint multiple-SNP analysis of GWAS summary statistics identifies additional variants influencing complex traits. Nature genetics 44, 369–375 (2012).

36. Chang, C.C. et al. Second-generation PLINK: rising to the challenge of larger and richer datasets. Gigascience 4, s13742-015-0047-8 (2015).

