## Supplementary Figure 1-2 for "Transcriptional and translational genetic control in blood contributes significantly to complex trait heritability and polygenic prediction"

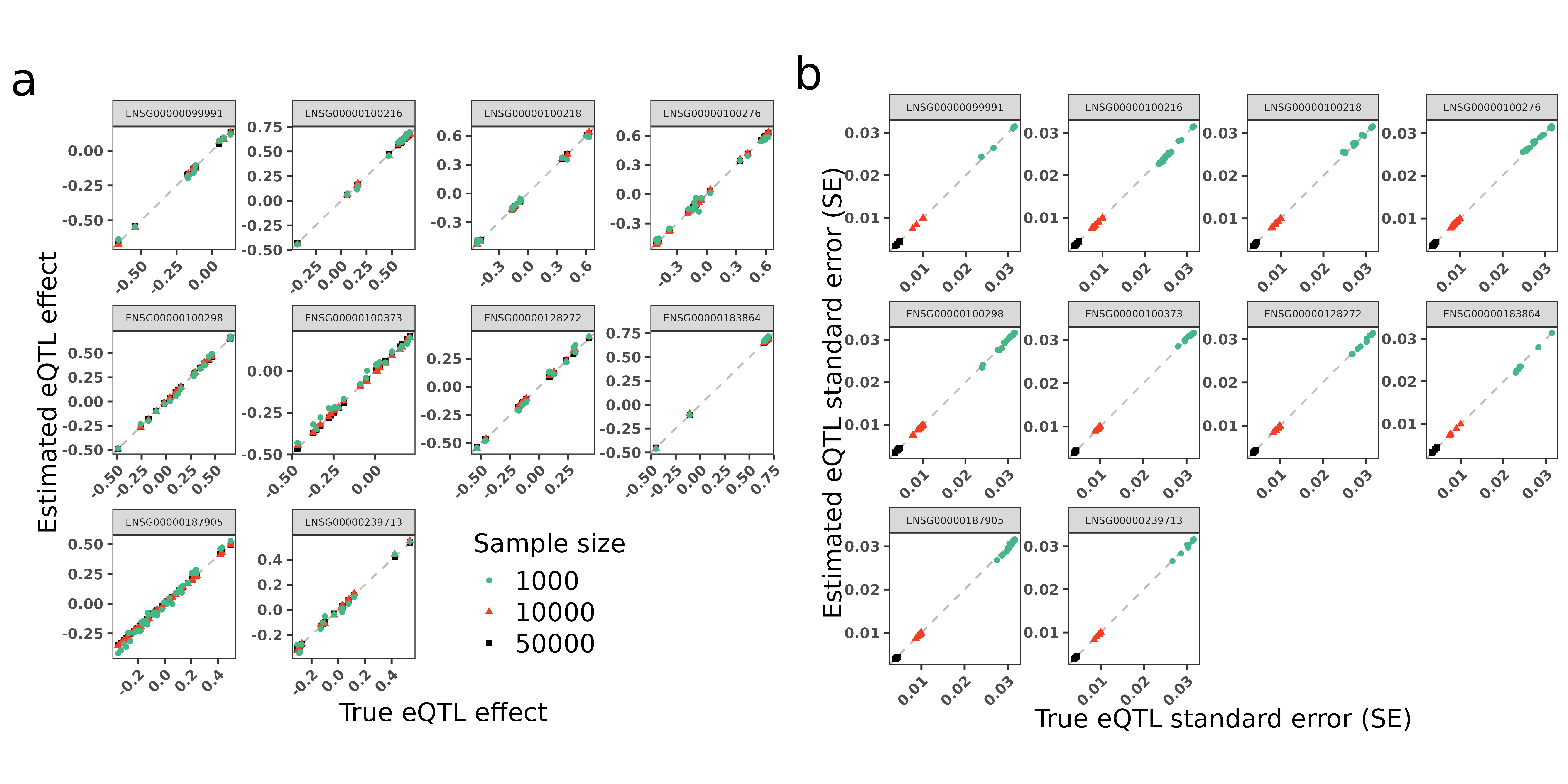

**Supplementary Figure 1 Performance of the down-sampling strategy under a 10-gene simulation.** a) Comparison of the estimated and true eQTL effect sizes under different sample sizes. b) Comparison of the estimated and true eQTL standard errors (SE) under different sample sizes.

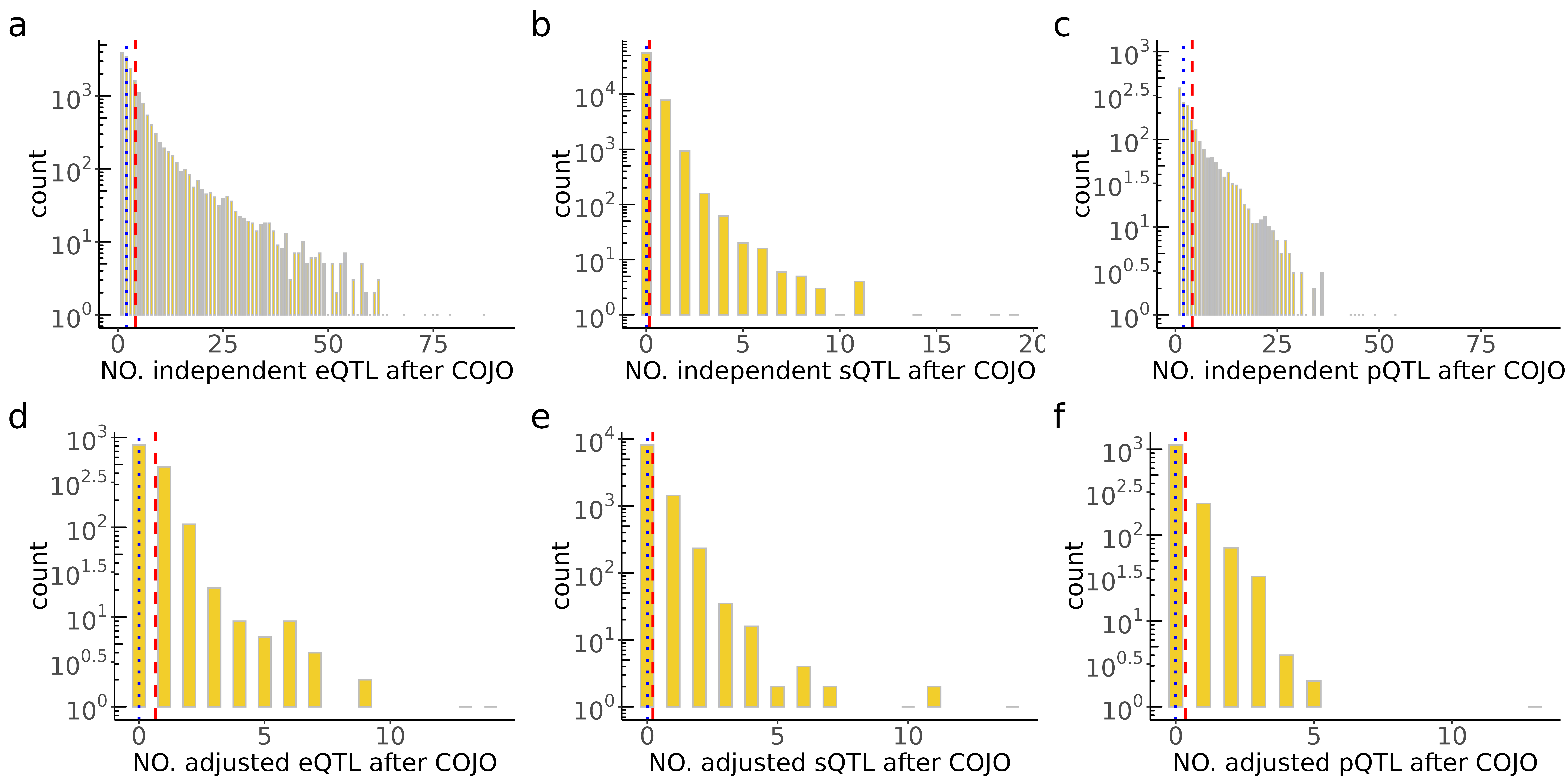

**Supplementary Figure 2 The distribution of different independent molQTLs after COJO analysis.** a) eQTL, b) sQTL, and c) pQTL with original sample sizes, d) adjusted eQTL, e) sQTL, and f) adjusted-sampled pQTL in common genes. The blue and red lines denote the mean and median, respectively.
